# Immunotherapy Significantly Improves Merkel Cell Carcinoma-Specific Survival: A Single-Cohort Propensity Score-Matched Analysis

**DOI:** 10.64898/2026.03.05.26347615

**Authors:** Sophia Z. Shalhout, Angela Fragano, Gabriella Chefitz, Tom W. Andrew, Kristina Lachance, Rima Kulikauskas, Paul Nghiem, Isaac Brownell

**Affiliations:** Division of Surgical Oncology, Department of Otolaryngology-Head and Neck Surgery, Mike Toth Head and Neck Cancer Research Center, Mass Eye and Ear, Boston, Massachusetts; Department of Otolaryngology-Head and Neck Surgery, Harvard Medical School, Boston, Massachusetts; Dermatology Branch, National Institute of Arthritis Musculoskeletal and Skin Diseases (NIAMS), National Institutes of Health (NIH), Bethesda, Maryland, USA; Translation and Clinical Research Institute, Newcastle University, Newcastle upon Tyne, UK; Department of Plastic and Reconstructive Surgery, Royal Victoria Infirmary, Newcastle Upon Tyne Hospital NHS Foundation Trust (NuTH), Newcastle upon Tyne, UK; Department of Dermatology, University of Washington, Seattle, WA, USA; Fred Hutch Cancer Center, Seattle, Washington, USA

**Keywords:** Merkel Cell Carcinoma, Immunotherapy, Immune checkpoint inhibitor, Disease-specific survival, Propensity score matching, Advanced skin cancer

## Abstract

**Background:** Immune checkpoint inhibitors (ICI) have improved outcomes in Merkel cell carcinoma (MCC). Population analyses suggest improved survival following the 2017 approval of ICI, but registry data lack treatment-level information including type of systemic therapy and initiation timepoint to directly estimate the benefit attributable to immunotherapy. This study compared Merkel Cell Carcinoma-specific survival between patients treated with first-line ICI versus cytotoxic chemotherapy.

**Methods:** Patients were identified from the Seattle Merkel Cell Carcinoma Registry. Among 1,517 patients with MCC, 463 received first-line systemic therapy with either ICI or chemotherapy. Propensity scores were estimated using logistic regression including AJCC 8th stage, age, sex, MCPyV status, and immunosuppression. One-to-one nearest-neighbor matching produced balanced cohorts of 133 ICI-treated and 133 chemotherapy-treated patients. Merkel Cell Carcinoma-specific survival from therapy initiation was analyzed using Kaplan-Meier and Cox proportional hazards models with follow-up administratively censored at five years.

**Results:** Baseline clinical characteristics were comparable between matched cohorts. ICI therapy was associated with significantly improved Merkel Cell Carcinoma-specific survival compared with chemotherapy (log-rank p<0.0001). Five-year Merkel Cell Carcinoma-specific survival was 56.8% (95% CI 46.8-65.6) for ICI versus 23.9% (95% CI 16.9-31.6) for chemotherapy. In multivariable stage-stratified Cox analysis, ICI remained independently associated with improved Merkel Cell Carcinoma-specific survival (HR 0.32, 95% CI 0.21-0.50; p<0.0001), while immunosuppression was associated with worse Merkel Cell Carcinoma-specific survival (HR 2.03, 95% CI 1.10-3.74; p=0.0228).

**Conclusions:** ICI therapy was associated with substantially improved MCC-specific survival compared with chemotherapy.

## Introduction

Immune checkpoint inhibitors (ICI) have reshaped outcomes in Merkel cell carcinoma (MCC). A recent analysis of the Surveillance, Epidemiology, and End Results (SEER) registry found improved 2-year relative survival following the 2017 approval of ICI for MCC.^1^ However, SEER lacks specific therapy data, preventing a precise estimation of the benefit attributable to ICI. We conducted a single-cohort, propensity score-matched (PSM) survival analysis to directly compare first-line ICI versus chemotherapy in patients with MCC.

## Methods

We queried the Seattle Merkel Cell Carcinoma Registry. Among 1,517 patients with MCC (1998-2024), 463 received either first-line ICI or chemotherapy. Propensity-scores were estimated using logistic regression including AJCC 8th stage, age, sex, MCPyV status, and immunosuppression (**Supplementary Methods**). One-to-one nearest-neighbor matching with a caliper of 0.2 yielded 133 ICI-treated and 133 chemotherapy-treated patients. Disease-specific survival (DSS), measured from initiation of first-line systemic therapy to MCC-specific death (or censored otherwise), was analyzed using Kaplan-Meier and Cox proportional-hazards models. Follow-up was administratively censored at five years to estimate fixed-horizon survival and ensure comparable follow-up. Number of covariates in the stage-stratified multivariable cox models were determined by events-per-parameter thresholds to minimize overfitting.

## Results

The distribution of age, sex, MCC tumor site, immunosuppression, MCPyV status, and stage at diagnosis was comparable between the PSM cohorts (**Table 1**). ICI therapy was associated with significantly improved DSS compared with chemotherapy (p<0.0001; **Figure 1**). Five-year DSS was higher in patients treated with ICI versus chemotherapy (56.8% [95%CI: 46.8%-65.6%] vs 23.9% [95%CI: 16.9%-31.6%]; log-rank p<0.0001).

**Table 1:** Characteristics for Propensity Score Matched Cohort of Patients Treated with Immune Checkpoint Inhibitors or Chemotherapy for Merkel Cell Carcinoma.

| Patient Characteristics | Propensity Score Matched Cohort |  |  |
| --- | --- | --- | --- |
|  | ICI<br>(N= 133) | Chemotherapy<br>(N= 133) | p-value |
| **Age** |  |  | 0.10 |
| Median age, years | 70 (18-93) | 67 (41-95) |  |
| **Race - no. (%)** |  |  | **0.05** |
| Black | 1 (0.8%) | 0 (0.0%) |  |
| Asian | 0 (0.0%) | 2 (1.5%) |  |
| White | 122 (92%) | 110 (83%) |  |
| Unknown | 10 (7.5%) | 19 (14.3%) |  |
| More than one race | 0 (0.0%) | 1 (0.8%) |  |
| Other | 0 (0.0%) | 1 (0.8%) |  |
| **Sex - no. (%)** |  |  | 0.38 |
| Male | 99 (74.4%) | 106 (79.7%) |  |
| **Primary Site of MCC - no. (%)** |  |  | 0.84 |
| Head/Neck | 37 (27.8%) | 36 (27.1%) |  |
| Extremities | 49 (36.8%) | 55 (41.4%) |  |
| Trunk | 19 (14.3%) | 15 (11.3%) |  |
| Unknown Primary | 28 (21.1%) | 27 (20.3%) |  |
| **Systemic Therapy - no. (%)** |  |  |  |
| Pembrolizumab | 78 (58.6%) | 0 (0.0%) | — |
| Nivolumab | 12 (9.0%) | 0 (0.0%) |  |
| Ipilimumab + Nivolumab | 1 (0.8%) | 0 (0.0%) |  |
| Avelumab | 40 (30.1%) | 0 (0.0%) |  |
| Cemiplimab | 2 (1.5%) | 0 (0.0%) |  |
| Cytotoxic Chemotherapy* | 0 (0.0%) | 133 (100%) |  |
| **Immunosuppression\*\* - no. (%)** |  |  | 0.75 |
| Yes | 23 (17.3%) | 26 (19.5%) |  |
| No | 110 (82.7%) | 107 (80.5%) |  |
| **MCPyV Status - no. (%)** |  |  | 0.41 |
| Positive | 67 (50.4%) | 61 (45.9%) |  |
| **Stage (AJCC 8<sup>th</sup> ed) - no. (%)** |  |  | 1.0 |
| Stage I | 23 (17.3%) | 23 (17.3%) |  |
| Stage II | 17 (12.8%) | 17 (12.8%) |  |
| Stage III | 75 (56.4%) | 75 (56.4%) |  |
| Stage IV | 16 (12.0%) | 16 (12.0%) |  |
| Unknown | 2 (1.5%) | 2 (1.5%) |  |

\*Cytotoxic Chemotherapy including regimens of carboplatin, cisplatin, etoposide, topotecan, irinotecan, and paclitaxel.
\*\*Immunosuppression including hematologic malignancies, solid organ transplant, autoimmune disease, HIV/AIDS.

**Figure 1:**
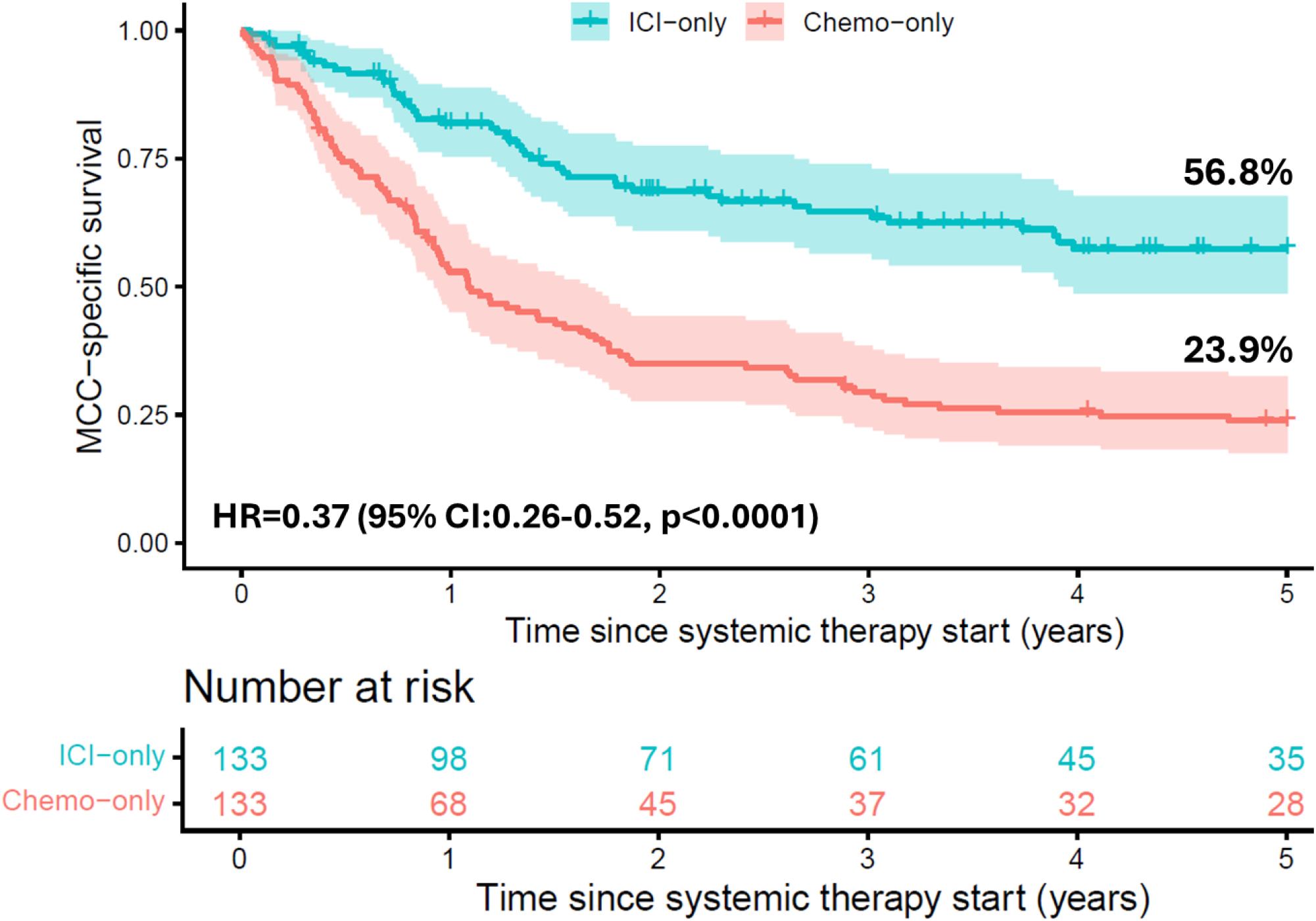
Immune Checkpoint Inhibitors Improve MCC-Specific Survival versus Chemotherapy. Kaplan-Meier curves for MCC-specific survival in patients treated with first-line immune checkpoint inhibitors (ICI, blue) or chemotherapy (red) in the propensity score-matched cohort, with follow up truncated at 5 years. Compared with chemotherapy, ICI therapy was associated with a significantly lower risk of MCC-specific death (log-rank p<0.0001). Five-year DSS was 2.4x higher in patients treated with ICI (56.8% [95%CI, 46.8%-65.6%]) versus chemotherapy (23.9% [95%CI, 16.9%-31.6%]; log-rank p<0.0001). Univariable and stage-stratified Multivariable Cox regression also found improved DSS associated with ICI treatment (HR=0.37; 95%CI, 0.26-0.52; p<0.0001; HR=0.32; 95%CI, 0.21-0.50; p<0.0001, respectively).

In a univariable Cox proportional hazards analysis (**Supplementary Table 1**), immunosuppression was associated with worse DSS (HR=1.98, 95%CI: 1.35-2.89; p<0.001), whereas ICI therapy was associated with improved DSS compared with chemotherapy (HR=0.37, 95%CI: 0.26-0.52; p<0.0001).

A stage-stratified Multivariable Cox regression model confirmed that treatment modality was independently associated with DSS after adjusting for clinically relevant variables (**Supplementary Figure 1**). ICI remained associated with improved DSS compared with chemotherapy (HR=0.32, 95%CI: 0.21-0.50; p<0.0001) among patients with the same stage, while immunosuppression remained associated with worse DSS (HR=2.03, 95%CI: 1.10-3.74; p=0.0228). No significant associations were observed for other variables after adjustment.

## Discussion

This case-level matched comparison demonstrates that first-line ICI therapy significantly improves MCC-specific survival compared to chemotherapy. Prior to the advent of ICI, systemic therapy for MCC yielded short-lived responses.^2^ The DSS benefit observed here is consistent with clinical trial data demonstrating durable disease control with ICI.^3^ These results also align with the long-term survival benefit seen in melanoma treated with ICI and population-level improvement observed in immunotherapy-era MCC survival.^1,4^ Immunosuppression remained an independent adverse prognostic factor, consistent with studies demonstrating attenuated ICI response and survival among immunocompromised patients with MCC.^5^ This study is limited by its retrospective single-cohort design and the potential for residual confounding by practice changes over time, despite propensity score matching and multivariable adjustment. Moreover, immunosuppression was analyzed as a composite variable, potentially masking etiologic heterogeneity.

## Conclusion

ICI therapy was associated with improved DSS compared with cytotoxic chemotherapy, with a 63% lower risk of MCC death and a 2.4-fold increase in 5-year survival.

## Data Availability

All data produced in the present study are available upon reasonable request to the authors contingent on appropriate data usage agreements and IRB approvals.

## Acknowledgment

This work was conducted in collaboration with The Merkel Cell Carcinoma Collaborative (MC3) Institute based at UW Medicine and Fred Hutch Cancer Center.

