## Supplemental Tables, Figures and Methods for "Immunotherapy Significantly Improves Merkel Cell Carcinoma-Specific Survival: A Single-Cohort Propensity Score-Matched Analysis"

**Mendeley Supplementary Materials:**

**Mendeley Supplementary Table 1: MCC-Specific Survival Univariable Cox Proportional Hazards Analysis in a Propensity-score Matched Cohort of Patients with Advanced Merkel Cell Carcinoma**

| Patient Characteristics | Total Cohort (N=266) | |
|---|---|---|
| | **HR (95% CI)** | **p-value** (bold < 0.05) |
| **ICI versus Chemotherapy** | 0.37 (0.26-0.52) | **<0.0001** |
| **Age** | 0.99 (0.98-1.01) | 0.539 |
| **Race** | - | global Wald p = 0.09 |
| White | Reference | - |
| Black | 2.01 (0.28-14.46) | 0.486 |
| Asian | 1.06 (0.15-7.55) | 0.958 |
| Other/Unknown | 1.75 (1.12-2.73) | 0.014 |
| **Sex** | | |
| Female | Reference | - |
| Male | 0.97 (0.66-1.43) | 0.883 |
| **Primary Site** | | |
| Head/Neck | Reference | - |
| Extremities | 1.01 (0.67-1.52) | 0.959 |
| Trunk | 1.25 (0.75-2.11) | 0.392 |
| Unknown Primary | 0.88 (0.54-1.41) | 0.583 |
| **Immunosuppression** | | |
| No | Reference | **-** |
| Yes | 1.98 (1.35-2.89) | **<0.001** |
| **Lymphovascular invasion** | | |
| No | Reference | - |
| Positive | 0.90 (0.63-1.29) | 0.566 |
| **MCPyV Status** | | |
| Not Positive | Reference | - |
| Positive | 0.86 (0.62-1.18) | 0.344 |
| **Sentinel Lymph Node Biopsy** | | |
| No | Reference | - |
| Yes | 0.83 (0.59-1.15) | 0.263 |
| **Sentinel Lymph Node Biopsy Results** | | |
| Negative | Reference | - |
| Positive | 1.05 (0.72-1.54) | 0.795 |
| **Surgical Excision** | | |
| No | Reference | - |
| Yes | 1.14 (0.81-1.61) | 0.458 |
| **Radiotherapy** | | |
| No | Reference | - |
| Yes | 1.17 (0.81-1.71) | 0.403 |
| **Stage (AJCC 8$^{th}$ ed) at Diagnosis** | | |
| Stage I | Reference | - |
| Stage II | 1.46 (0.78-2.75) | 0.236 |
| Stage III | 1.51 (0.94-2.43) | 0.090 |
| Stage IV | 1.47 (0.78-2.75) | 0.233 |
| Unknown | 1.86 (0.44-7.95) | 0.403 |

**<u>Mendeley Supplementary Figure 1</u>: Multivariable Stage-Stratified Cox Proportional Hazards Model for MCC-Specific Survival in a Propensity Score Matched Cohort of Advanced Merkel Cell Carcinoma**

Forest plot displays adjusted hazard ratios (HRs) and 95% confidence intervals (CIs) for MCC-specific survival in the propensity score matched cohort. Multivariable Cox proportional hazards modeling included treatment [immune checkpoint inhibitor (ICI) vs chemotherapy], age, sex, primary site, MCPyV status, and immunosuppression. Covariates were selected *a priori* based on established clinical relevance to control for potential confounding, while maintaining appropriate events-per-parameter thresholds to support stable multivariable estimation and avoid overfitting. Disease stage was incorporated via stratification in the Cox model, allowing separate baseline hazard functions for each Stage I, II, III, IV, and Unknown without estimating stage specific hazard ratios. Reported hazard ratios therefore represent associations conditional on stage, reflecting comparisons among patients within the same stage category. Robust sandwich standard errors were estimated with clustering by matched set to account for the matched design. Among patients with the same stage, ICI therapy was independently associated with improved MCC-specific survival compared with chemotherapy (HR=0.32, 95%CI 0.21-0.50; p<0.0001), whereas immunosuppression was associated with worse survival (HR=2.03, 95%CI 1.10-3.74; p=0.0228). Age, sex, primary site, and MCPyV status were not statistically significant after stage stratification and multivariable adjustment. Hazard ratios are displayed on a logarithmic scale; points represent point estimates and horizontal bars denote 95% CIs. The dashed vertical line indicates HR=1.0. Global likelihood ratio, Wald, and robust score tests were statistically significant (p<0.0001), with a C-index of 0.70 (95%CI 0.62-0.74).

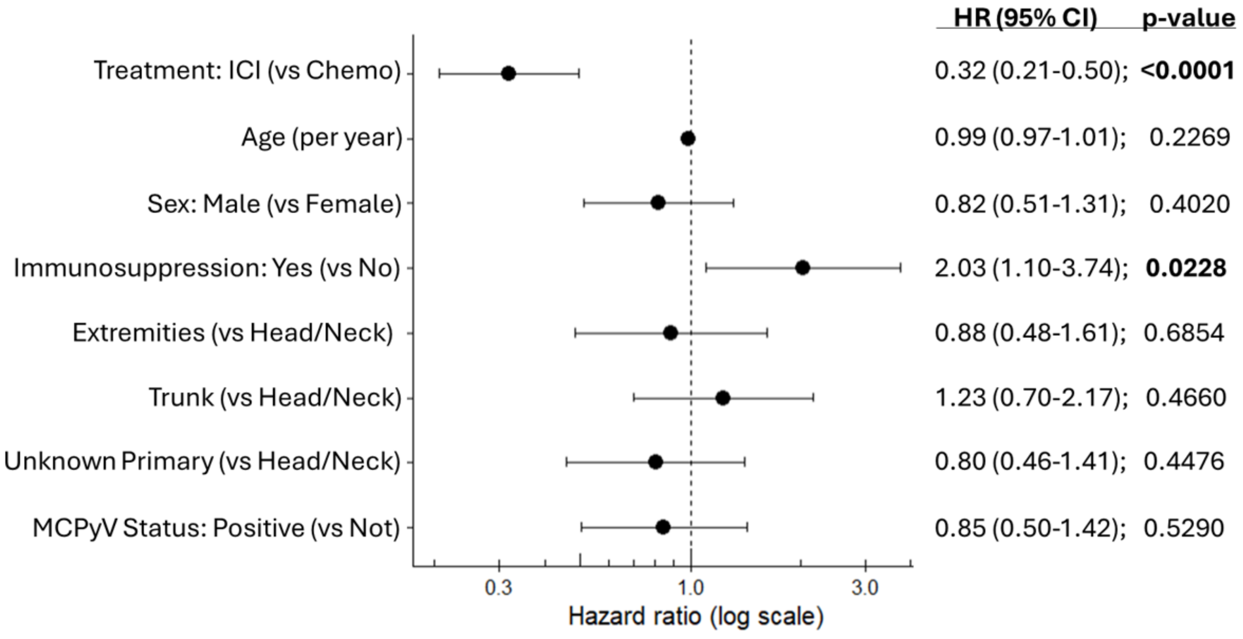

**Mendeley Supplementary Methods:**

*Study Cohort*

We analyzed data from the Seattle Merkel Cell Carcinoma (MCC) Registry, a prospectively maintained longitudinal cohort. Among 1,517 patients with pathologically confirmed MCC, 463 who received first-line systemic therapy with immune checkpoint inhibitors (ICI) or cytotoxic chemotherapy between 1998 and 2024 were eligible. Patients with incomplete treatment start dates or missing outcome data were excluded. Patients who received concurrent immune checkpoint inhibitor therapy and cytotoxic chemotherapy as first-line systemic treatment were excluded to preserve mutually exclusive treatment groups.

*Exposure and Covariates*

First-line ICI included treatment with Pembrolizumab, Nivolumab, Ipilimumab/Nivolumab, Avelumab or Cemiplimab. First-line cytotoxic chemotherapy was defined as treatment with conventional, non-immunologic antineoplastic agents. Regimens included platinum-based combinations, most commonly carboplatin or cisplatin administered with etoposide (including VP-16), as well as other cytotoxic agents used alone or in combination, including topotecan, irinotecan, and paclitaxel (taxol). Platinum-etoposide combinations constituted the predominant first-line cytotoxic regimen in this cohort.

Baseline covariates were specified *a priori* based on clinical relevance and included age, sex, AJCC 8th edition stage, tumor Merkel cell polyomavirus (MCPyV) status, and immunosuppression. Immunosuppression was a composite category defined as hematologic malignancy, autoimmune disease requiring systemic immunosuppression, solid organ transplantation requiring immunosuppressive therapy, and/or HIV/AIDS.

*Propensity Score Matching*

Propensity scores were estimated using multivariable logistic regression to model the probability of receiving immune checkpoint inhibitor (ICI) versus chemotherapy as first-line systemic treatment. Covariates included in the propensity score model were selected *a priori* based on clinical relevance and prior literature and comprised age at diagnosis, sex, AJCC 8th edition stage at diagnosis, tumor Merkel cell polyomavirus (MCPyV) status, and immunosuppression status. For matching covariates with missing data, an "unknown/not reported" category was included to retain patients in the propensity score estimation and matching process.

One-to-one nearest-neighbor matching without replacement was performed using a caliper width of 0.2 on the logit of the propensity score to reduce poor-quality matches and residual confounding. Patients outside the region of common support were excluded. This approach yielded a well-matched cohort of 266 patients, consisting of 133 patients treated with ICI in the first line and 133 patients treated with chemotherapy in the first line. Covariate balance before and after matching was evaluated using standardized mean differences (SMDs), with values less than 0.1 considered indicative of adequate balance. Balance diagnostics were also evaluated visually using Love plots to compare the distribution of covariates across treatment groups before and after matching. The matched cohort was used for all subsequent survival analyses.

*Outcome and Survival Analysis*

Disease-specific survival (DSS) was defined as the time from initiation of first-line systemic therapy (ICI or cytotoxic chemotherapy) to MCC-specific death. Patients were censored at non-MCC death or last follow-up, with follow-up administratively censored at five years to estimate fixed-horizon survival and ensure comparable follow-up duration. DSS was estimated using Kaplan-Meier methods and differences between treatment groups assessed using the log-rank test. Multivariable Cox proportional hazards regression models were then constructed within the propensity score-matched cohort to estimate hazard ratios (HRs) and 95% confidence intervals (CIs) for associations between clinical covariates and DSS. Stage at diagnosis was incorporated through stratification in the Cox model, allowing separate baseline hazard functions by stage category without estimating stage-specific hazard ratios. Robust sandwich standard errors clustered by matched set were used to account for the matched design.

*Multivariable Cox Regression and Robust Variance*

To evaluate the independent association between treatment modality and MCC-specific survival within the propensity score matched cohort, a multivariable Cox proportional hazards regression model was constructed. The outcome was time to MCC-specific death within five years of initiation of first-line systemic therapy with censoring at non-MCC death, last follow-up, or five years, whichever occurred first.

The primary exposure of interest was treatment modality, defined as immune checkpoint inhibitor therapy versus chemotherapy. Prespecified covariates were selected based on established clinical relevance and included age at diagnosis (modeled continuously), sex, primary MCC tumor site, immunosuppression status, and Merkel cell polyomavirus status. Inclusion of these variables allowed control for residual confounding after matching and provided a doubly robust estimation framework. Model complexity was limited according to conventional events-per-parameter considerations to reduce the risk of overfitting and unstable coefficient estimates. The final model satisfied accepted thresholds for stable multivariable Cox estimation. Analyses were conducted using a complete case approach; individuals with missing covariate data were excluded from the multivariable Cox model.

Disease stage (Stage I–IV and Unknown) was incorporated via stratification, allowing separate nonparametric baseline hazard functions for each stage category without estimating stage-specific hazard ratios. Treatment and covariate effects therefore represent associations conditional on stage, reflecting comparisons among patients within the same stage stratum.

Because propensity score matching induces correlation within matched sets, standard errors and confidence intervals were estimated using a robust sandwich variance estimator clustered by matched set. Global model inference was evaluated using likelihood ratio, Wald, and robust score tests. Discriminative performance was quantified using the concordance index (C-index).

*Statistical Software*

All analyses were conducted using R (version 4.5.0). No formal adjustment for multiple comparisons, including Bonferroni correction, was applied, as analyses were prespecified and hypothesis-driven, with treatment effect estimates interpreted in the context of effect sizes and confidence intervals rather than isolated p-values.
